# Determinants of Short Birth Intervals among married women in Karachi, Pakistan

**DOI:** 10.1101/2020.08.13.20174110

**Authors:** Sidrah Nausheen, Maria Bhura, Kristy Hackett, Imtiaz Hussain, Zainab Shaikh, Arjumand Rizvi, Uzair Ansari, David Canning, Iqbal Shah, Sajid Bashir Soofi

## Abstract

**Introduction:** Birth spacing is a critical pathway to improving reproductive health. The World Health Organization recommends a minimum of 33-month interval between two consecutive births to reduce maternal, perinatal, and infant morbidity and mortality. Our study evaluated factors associated with short birth intervals (SBIs) of less than 33 months between two consecutive births, in three peri-urban municipalities in Karachi, Pakistan.

**Methods:** We used data from a cross-sectional study among married women of reproductive age (MWRA) who had at least one live birth in the six years preceding the survey (N=2394). Information regarding their sociodemographic characteristics, reproductive history, fertility preferences, family planning history, and a six-year reproductive calendar were collected. To identify factors associated with SBIs, we fitted simple and multiple Cox-proportional hazards models and computed hazard ratios (HR) with their 95% confidence intervals (CI).

**Results:** The median birth interval was 25 months (IQR: 14-39 months), with 22.9% of births occurring within 33 months of the index birth. Women’s increasing age [25-29 years (aHR=0.64, 95% CI: 0.54-0.77), 30+ years (aHR=0.30, 95% CI: 0.23-0.40) compared to <25 years]; secondary education [aHR 0.78. 95% CI: 0.65-0.93], intermediate education [aHR 0.63, 95% CI: 0.49-0.82], higher education (aHR=0.71, 95% CI: 0.53-0.96) compared to no education, and a male child of the index birth (aHR=0.79, 95% CI: 0.68-0.92) reduced the likelihood of SBIs. Women’s younger age <20 years [aHR 1.32, 95% CI 1.03-1.70] compared to 20-24 years, and those who did not use contraception within 9-months of the index birth had a higher likelihood for SBIs for succeeding birth compared to those who used contraception (aHR=2.33, 95% CI: 2.01-2.70).

**Conclusion:** This study evaluates factors associated with birth spacing practices among married women of childbearing age in urban settlements of Karachi. Our study shows that birth intervals in the study population are lower than the national average. To optimize birth intervals, programs should target child spacing strategies and counsel MWRA on the benefits of optimal birth spacing, family planning services and contraceptive utilization.

**Ethics:** The study received ethical approval from the Ethical Review Committee (ERC) at the Aga Khan University (AKU) (4964-Ped-ERC-17) and the Institutional Review Board (IRB) at the Harvard T.H. Chan School of Public Health (IRB17-1864). Informed written consent was obtained from each study participant. Women who were unable to sign provided consent with a thumb impression in the presence of witnesses.

**Strengths and limitation:**

1. This is the first study that has investigated birth spacing in urban areas of Karachi, Pakistan
2. It is a cross sectional study that has employed a three-stage random sampling design i.e. at cluster level, at household level, and at individual level.
3. Participants were selected from low-income areas of Karachi, therefore not representative of metropolitan population of Karachi.
4. There may be an underrepresentation of birth intervals because the study did not consider abortions or miscarriages.
5. The study only considered births in six-year calendar time and therefore births occurred before or after this calendar time were considered as no-event.
6. There may be underestimation in birth intervals where women did not give birth since her last-born in this six-year calendar time.

## Background

Birth spacing is integral to improving reproductive health. The World Health Organization (WHO) recommends a minimum 24-month birth-to-pregnancy interval, or a 33-month interval between two consecutive births to reduce the risk of adverse maternal, perinatal, and infant health outcomes.^1^ Birth spacing is highly influenced by socioeconomic, demographic, cultural, and behavioural characteristics.^2^ Short birth-to-birth intervals, also known as, short birth intervals (SBIs) are associated with poor neonatal and infant outcomes,^3^ including low birth-weight,^4^ preterm births,^5^ small-for-gestational-age,^6^ neonatal mortality,^7,8^ and infant mortality.^4,9,10^ Short birth-to-pregnancy intervals are also associated with a 61% increased risk in neonatal mortality and a 48% increased risk in under-5 mortality if the interval is less than 24 months.^11^

Similarly, maternal health is negatively impacted by SBIs, where women do not have sufficient time to physically recuperate from their previous pregnancy.^12^ Closely spaced pregnancies increase maternal nutrition depletion, resulting in a reduction of the mother’s nutritional status.^13^ Birth-to-pregnancy intervals of less than six months can significantly increase the odds of maternal mortality by 150% (95% CI 22-438%), and are associated with an increased risk of third trimester bleeding, premature rupture of membranes, postpartum endometriosis and anaemia.^14^ A systematic review of studies from Ethiopia found that women with birth-to-pregnancy intervals of less than two years were twice at risk of developing anaemia during their next pregnancy since repeated pregnancies tend to deplete a woman’s iron stores.^15^ However, systematic reviews have reported conflicting and low-quality evidence between maternal health outcomes and SBIs.^16,17^

Longitudinal data on singleton live births in Bangladesh found that shorter intervals between birth and pregnancy were associated with higher infant and child mortality, and longer birth intervals improved child survival.^18,19^ Several studies have found associations between SBIs and neonatal and infant mortality in both low-and middle-income and high-income countries over time.^20-25^ Systematic reviews and Demographic Health Survey (DHS) analyses have also studied the impact of SBIs on infant mortality, particularly in low-income countries.^26,27^ SBIs are associated with infant morbidity and poor health outcomes in multiple ways, for both the older child as well as the one born after the SBI. Women with closely spaced pregnancy may less likely to attend antenatal care services (which are critical for monitoring pregnancy and identifying complications) because they have other child to take care of.^28^ Furthermore, lactation may be impaired due to maternal nutritional depletion and they may be unable to provide adequate breastfeeding to their older infant.^28^ Children who are closely spaced are more likely to compete for resources, such as breastmilk, parental attention, and time.^24,29^

Pakistan has a population of over 216.6 million people in 2019 and is currently the fifth most populous country in the world, with an annual population growth rate of 2.1% and a fertility rate of 3.6 children per woman in 2017.^30,31^ The country possesses a maternal mortality ratio of 276 deaths per 100,000 live births, neonatal mortality of 42 deaths per 1,000 live births, and infant mortality at 62 deaths per 1,000 live births.^32-34^ The median age at first birth is 22.8 years among MWRA. Moreover, the use of any method of family planning by currently married women is 34%, with 25% using a modern method and 9% using a traditional method of contraception.^35^ Although Pakistan’s median birth interval is 28.2 months according to Pakistan Demographic and Health Survey (PDHS) 2017-18, 37% of the births occur within 24 months of the preceding birth.^35^ This statistic is higher among younger women, where women aged 15-19 years have birth intervals which are 12.4 months shorter, on average, than women aged 30-39 years.^35^

An earlier study across 21 low and middle income countries (LMICs) revealed that Pakistan has one of the highest percentages (60%) of short birth-to-pregnancy intervals (<23 months after birth) with 31% unmet need for spacing and 29% unmet need for limiting.^36^ The unmet need for spacing and limiting pregnancies in Pakistan is 17%, indicating that several women who want to space or limit pregnancies do not use any method to do so.^35^ Therefore, opting for family planning and contraceptive use after childbirth can help women achieve healthy spacing of pregnancies.^37^ In Pakistan, preference for a male child is deeply entrenched, therefore couple’s wait before moving to subsequent pregnancy is short as long as desired number of son(s) are not born.^38,39^ A recent study from Pakistan has reported that birth intervals of less than 24 or 18 are higher among women without one or more sons.^39^ Other predictors that contribute to birth intervals in other studies include wealth indices, women’s education, maternal age, later start of reproductive years, gender of an index child, and parity according to studies conducted in Bangladesh, Iran, and Ethiopia.^40-42^ However, there is a lack of data on birth intervals in Pakistan. Our study seeks to explore the socioeconomic, demographic, and reproductive factors associated with SBIs of less than 33 months using retrospective data from urban populations in Karachi, Pakistan.

## Methods

### Study Design

This study draws on data from an evaluation of the Willows Program (https://proiects.iq.harvard.edu/willowsimpactevan. a community-based reproductive health program that provides family planning information, education, and referral through household visits to women of reproductive age (WRA). The parent study assessed the effect of the Willows program on modern contraceptive use with an aim to guide future programming for family planning interventions in Pakistan. The parent study was a retrospective cross-sectional assessment that took place between August and December 2018.

### Study setting and participants

This cross-sectional study was conducted in Korangi Town, PIB Colony, and Dalmia/Shanti Nagar, three peri-urban municipalities in Karachi, Pakistan. All areas are home to both locals and migrants from within the country, as well as Afghanistan, Bangladesh, and Burma, and have a majority of Muslim population. Women were eligible to participate in the study if they were married, usual household members, spoke at least one of the four commonly spoken languages (Urdu, Pushto, English, or Sindhi), were between the ages of 15-49 years, and self-reported themselves as fertile.

### Sample size and sampling strategy

For a parent study, a sample size of 1836 (~2000) from each area intervention and control area was required assuming an estimated modern contraceptive prevalence rate (mCPR) of below 30% in selected areas, methodology has been described in detail elsewhere. A three-stage random sampling design was carried out in STATA using a uniform [0,1] random number generator with a fixed seed. First, we used Geographical Information Systems (GIS) technology to construct a sampling frame with distinct area and cluster demarcation of the survey sites, forming 708 clusters in total. Next with a goal of an average of 60 households per enumeration area, we randomly selected 220 clusters, with 110 clusters from Korangi Town, and 110 clusters from PIB Colony and Dalmia/Shanti Nagar combined. Since PIB colony and Dalmia/Shanti Nagar are smaller in geographical and population size compared to Korangi town, therefore, equal numbers of clusters were selected from Korangi Town and PIB colony and Dalmia/Shanti Nagar combined. Proceeding that, an android application for household listing questionnaire was developed to assess the number of women between 15-49 years of age.. If more than one WRA lived in a selected household, we randomly selected one from the household.

### Data collection

We conducted face to face interviews with eligible women using a structured tablet-based questionnaire on the CommCare application. The survey questionnaire included a range of topics on women’s reproductive health, including information on socio-demographic characteristics of women and their husband, reproductive history, obstetric history, family planning history, fertility preferences, and a reproductive calendar of pregnancies, births, terminations and contraceptive use for the preceding six years. This study used a month-by-month calendar, similar to those collected in DHS and was based on a five-year recall period.

### Data analysis

#### Measures and outcomes

Information on birth intervals was analyzed using the contraceptive calendar for all participating women. Of all (4336) the randomly selected women, 4193 consented for participation in this retrospective survey. Of these, 2394 women who had given live birth at least once in the six years preceding the survey by using the calendar data were included in the analysis, and a total of 1799 MWRA were excluded because of no live birth in the five years preceding the survey or their pregnancies resulted in abortions or miscarriages. Index births were defined as the birth proceeding the birth interval. We assessed the association between birth spacing and sociodemographic characteristics, including woman’s age at index birth, woman’s education, husband’s education, wealth quintiles, ethnicity, sex of the infant, contraceptive uptake within nine months of the index birth, and length of the first birth interval in months. In examining the determinants of SBIs, we defined an event as the interval between the index birth and the next birth (live or stillborn) of less than 33 months, corresponding to recommendations by the WHO.^1^ Women who did not give birth until the end of the follow-up period were considered no-event by the survey as information only until the time of the interview was recorded.

#### Statistical analysis

Since this study considered time-to-event data, a survival analysis technique was carried out using Cox proportional hazards model. The Kaplan-Meier curve was used to estimate the median duration of the birth interval. Cox proportional hazard model was used to determine predictors of SBI. We initially performed bivariate analyses to examine the association between explanatory variables and the outcome variable (model A). A multivariable model was adjusted for all covariates (model B). Another multivariable Cox proportional hazard model was fitted by including variables with p<0.2 in the bivariate model (model C) using a backward elimination method, and variables with p<0.05 were retained within the model. Hazard Ratios (HRs) and their 95% confidence intervals (CIs) were computed with statistical significance determined at the 5% level (p<0.05). All analysis account clustering for the sampling design and women level using clustered-robust standard errors. The model was checked for multicollinearity using variance inflation factor using cutoffs of ≥10. All analyses were performed in STATA version 15.

### Patient and Public Involvement

The public was not involved in the design of the research tools, but they were part of the study. The key findings will be shared with their representatives as part of the dissemination plan at local level.

## Results

### Descriptive results

A total of 4336 MWRA were approached; of those 4193 women consented for participation in this retrospective survey. A total of 1799 MWRA were excluded because of no birth history, and 2394 women were included in our analysis, who had given birth to a total of 3641 children in the six years preceding the survey. Of the total births, 833 (22.9%) occurred in less than 33 months of the index birth; and the median birth interval in our study was 25 months (IQR: 14-39 months). Descriptive results for participants are presented in Table 1 with median and interquartile ranges for birth intervals in months for each category.

**Table 1:**
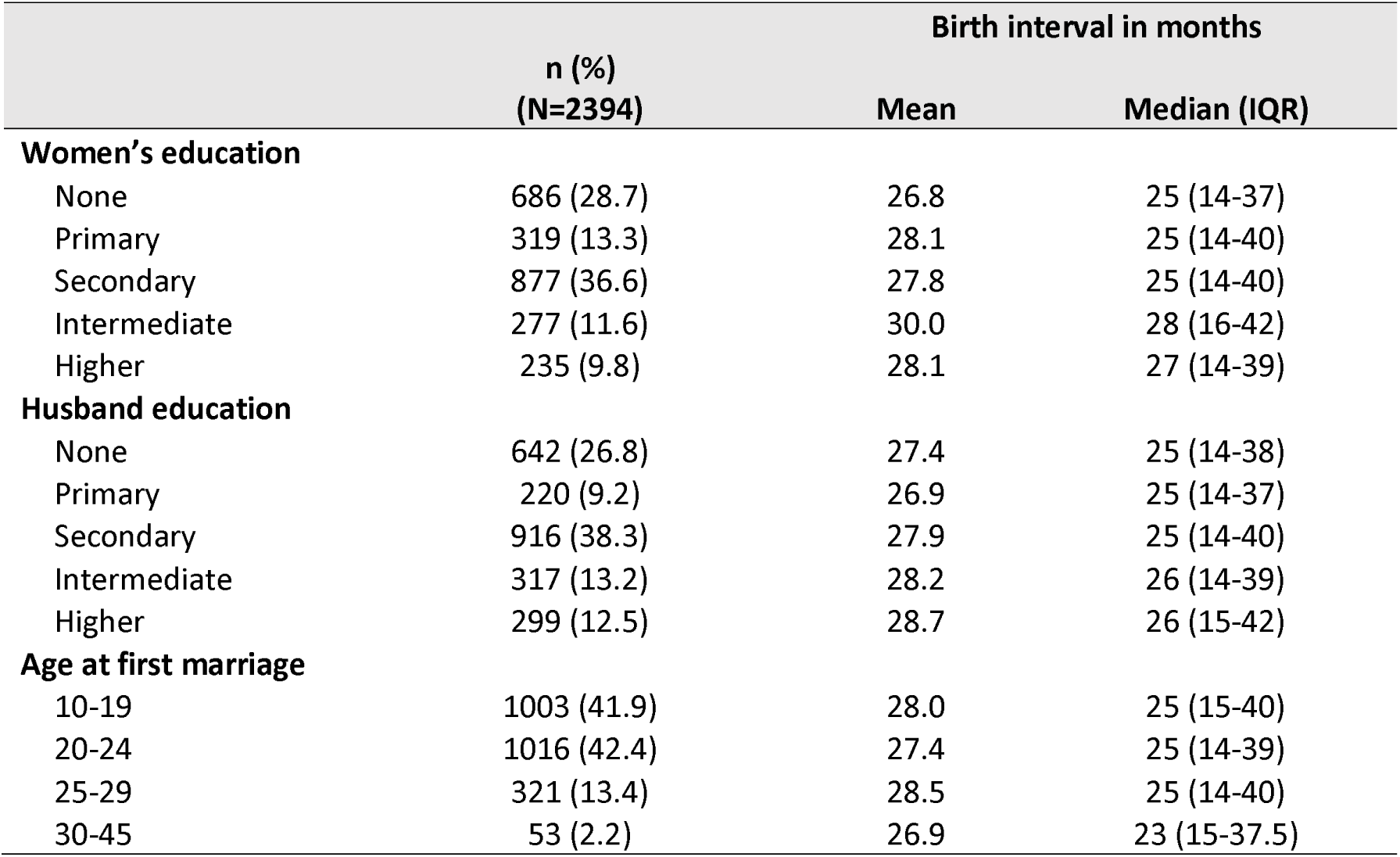
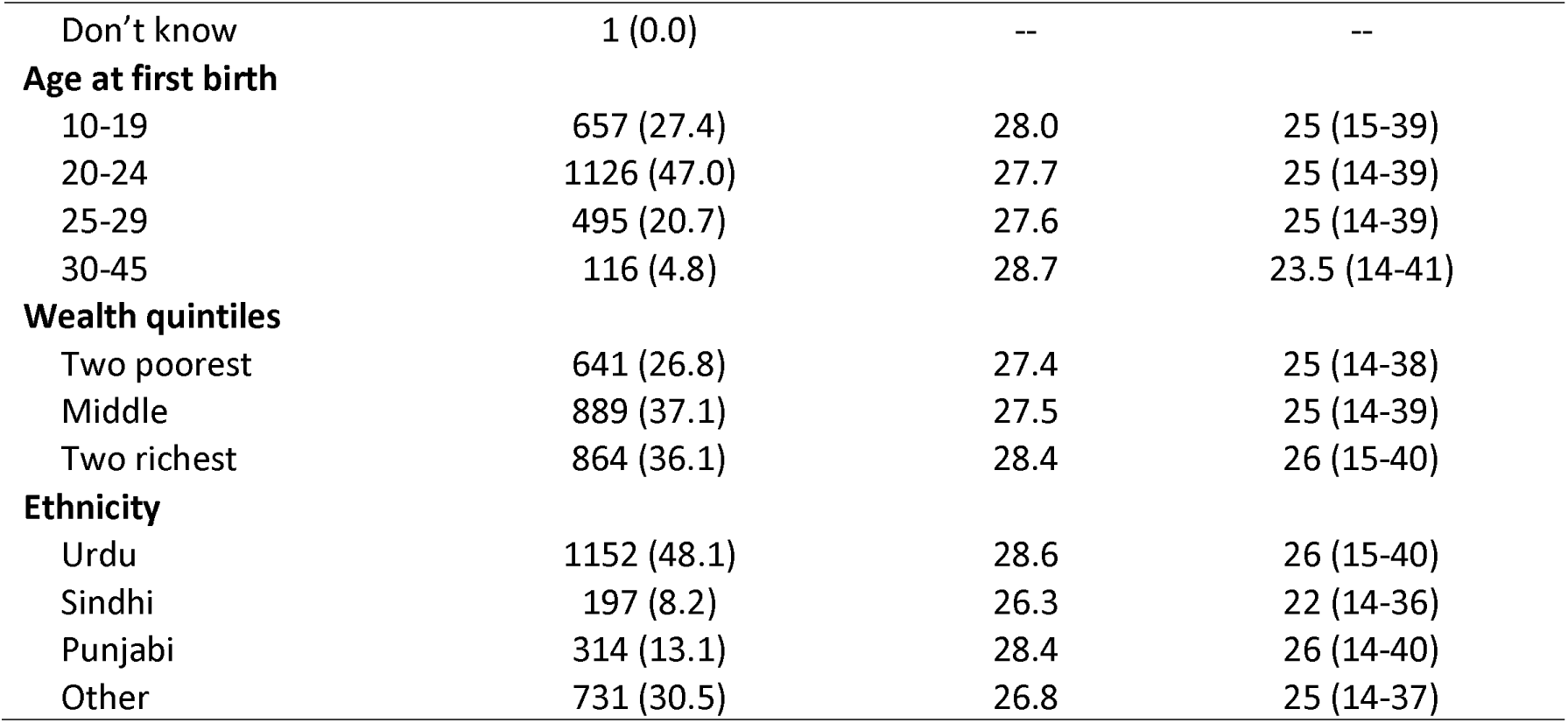
Percent distribution of socio-demographic characteristics of participants with mean and median birth interval (N=2394)

One in three women in our study had achieved secondary education (36.6%), with higher than secondary education being the least common (9.8%), and about one quarter (28.7%) women had no formal education. Similarly, one in three husbands had achieved secondary education (38.3%) and quarter of them had no education. Majority of our sample (84.3%) were married between 10-24 years of age, and 47.0% had their first birth between 20-24 years of age. About half the respondents (48.1%) belonged to an Urdu-speaking caste. From all index births included in our study, 32.5% were born when their mothers were 20-24 years of age, and 39.9% between 25-29 years of age. Majority of women belonged to middle wealth quintile (37.1%), while a similar number belonged to combined two richest quintiles (36.1%), and a quarter belonged to combined two poorest two quintiles (26.8%) (Table 2).

**Table 2:** Percent distribution of births with mean and median birth intervals (N=3641)

| Birth interval in months |
| --- |

|  | n (%)<br>N=3641 | Mean | Median (IQR) |
| --- | --- | --- | --- |
| **Total** |  | 27.8 | 25 (14-39) |
| **Age of woman at index birth** |  |  |  |
| <20 | 244 (6.7) | 27.4 | 25 (15.5-36.5) |
| 20-24 | 1182 (32.5) | 27.2 | 25 (15-37) |
| 25-30 | 1453 (39.9) | 27.1 | 25 (14-39) |
| >30 | 762 (20.9) | 29.9 | 28 (15-42) |
| **Contraceptive use within 9 months after index birth** |  |  |  |
| Used | 2612 (71.7) | 28.8 | 27 (15-41) |
| Did not use | 1029 (28.3) | 25.5 | 22 (14-35) |
| **Contraceptive methods used within 9-months after index birth** |  |  |  |
| Modern | 1791 (68.5) | 28.4 | 26 (14-41) |
| Traditional | 668 (25.6) | 29.8 | 28 (16-41) |
| Both | 153 (5.9) | 29.1 | 27 (16-40) |
| **Gender of index child ‡** |  |  |  |
| Male | 1732 (51.9) | 29.0 | 27 (15-41) |
| Female | 1603 (48.1) | 27.2 | 25 (14-38) |

‡ Denominator was 3335 for this variable as some of the children were the index birth.

When asked about contraceptive use within nine months of the index birth, about a quarter (28.3%) of participants did not use contraception (Table 2). Among those who used contraception, more than half (68.5%) used modern contraceptive methods, a quarter (25.6%) used traditional methods, and 5.9% used both modern and traditional methods. Women who did not use contraceptive methods had a shorter birth interval (median: 22 months, IQR: 14-35 months) than those who used modern contraceptive methods (median: 26 months, IQR: 14-41 months) or traditional contraceptive methods (median: 28 months, IQR: 16-41 months) (Table 2). Birth intervals varied slightly depending on the sex of the index birth. Data reveals that length of succeeding birth interval is shorter when the sex of index child is female, and this puts woman in even greater pressure to try for a male child earlier (Table 2).

### Predictors of short birth intervals

Bivariate analyses of predictors of SBIs (<33 months) are displayed in Table 3. They indicate that women aged 25-30 years and women who were greater than 30 years of age were less likely to have a SBIs compared to those younger than aged 20-24 years. However, the likelihood of SBI was higher among women less than 20 years old compared to women 20-24 years of age. Mothers who received secondary, intermediate, and higher education were also less likely to have SBIs than those who received no formal education. Likewise, husbands who received intermediate and higher were also less likely to have a SBI for the subsequent birth. Couples who did not use contraceptives within nine months were more likely to have SBIs. SBIs were also associated with the gender of the child born prior to the index birth; wealth quintiles, where those belonging to the middle and richest wealth quintiles were less likely to have SBIs, and ethnicity, with those belonging to a Sindhi or other background more likely to have SBIs compared to Urdu speaking families (Table 3).

**Table 3:** The Cox-Regression analysis of predictors of short birth interval (birth interval <33 months)

|  | Model A – Bivariate |  | Model B – Multivariate (all variables) |  | Model C – Multivariate (reduced) |  |
| --- | --- | --- | --- | --- | --- | --- |
|  | HR (95% CI) | P-value | Adjusted HR (95% CI) | P-value | Adjusted HR (95% CI) | P-value |
| **Woman age at index birth** |  |  |  |  |  |  |
| <20 | 1.29 (1.04-1.59) | 0.017 | 1.36 (1.05-1.76) | 0.02 | 1.32 (1.03-1.70) | 0.029 |
| 20-24 | 1 | . | 1 | . | 1 | . |
| 25-30 | 0.67 (0.58-0.78) | < 0.0001 | 0.61 (0.51-0.73) | < 0.0001 | 0.64 (0.54-0.77) | < 0.0001 |
| >30 | 0.38 (0.30-0.47) | < 0.0001 | 0.28 (0.21-0.38) | < 0.0001 | 0.30 (0.23-0.40) | < 0.0001 |
| **Mother education** |  |  |  |  |  |  |
| None | 1 | . | 1 | . | 1 | . |
| Primary | 0.87 (0.71-1.06) | 0.164 | 0.87(0.69-1.08) | 0.202 | 0.84 (0.68-1.05) | 0.132 |
| Secondary | 0.82 (0.7-0.95) | 0.010 | 0.79(0.65-0.96) | 0.020 | 0.78(0.65-0.93) | 0.005 |
| Intermediate | 0.62 (0.49-0.79) | < 0.0001 | 0.64(0.48-0.86) | 0.003 | 0.63(0.49-0.82) | 0.001 |
| Higher | 0.71 (0.55-0.92) | 0.009 | 0.71(0.50-1.02) | 0.061 | 0.71(0.53-0.96) | 0.026 |
| **Husband education** |  |  |  |  |  |  |
| None | 1 | . | 1 | . |  |  |
| Primary | 0.93 (0.73-1.19) | 0.580 | 0.95(0.73-1.23) | 0.695 |  |  |
| Secondary | 0.89 (0.77-1.04) | 0.163 | 0.99(0.83-1.19) | 0.929 |  |  |
| Intermediate | 0.79 (0.63-0.98) | 0.038 | 0.98(0.75-1.29) | 0.906 |  |  |
| Above | 0.69 (0.56-0.88) | 0.002 | 0.97(0.72-1.30) | 0.830 |  |  |
| **Contraceptive use within 9months after index birth** |  |  |  |  |  |  |
| Use | 1 | . | 1 | . | 1 | . |
| Did not use | 2.53 (2.2-2.91) | < 0.0001 | 2.28 (1.96-2.66) | < 0.0001 | 2.33(2.01-2.70) | < 0.0001 |
| **Age at first marriage** |  |  |  |  |  |  |
| 10-19 | 1 | . | 1 | . | 1 | . |
| 20-24 | 0.96 (0.84-1.11) | 0.562 | 1.07(0.85-1.35) | 0.548 | 1.24(1.05-1.48) | 0.013 |
| 25-29 | 0.86 (0.7-1.05) | 0.156 | 1.02(0.67-1.54) | 0.92 | 1.55(1.18-2.03) | 0.002 |
| 30-45 | 1.01 (0.61-1.63) | 0.994 | 1.64(0.71-3.81) | 0.247 | 2.79(1.59-4.90) | < 0.0001 |
| **Age at first birth** |  |  |  |  |  |  |
| 10-19 | 1 | . | 1 | . |  |  |
| 20-24 | 0.96 (0.83-1.12) | 0.602 | 1.17(0.93-1.49) | 0.181 |  |  |
| 25-29 | 0.96 (0.81-1.16) | 0.743 | 1.68(1.15-2.43) | 0.007 |  |  |
| 30-45 | 0.89 (0.62-1.26) | 0.521 | 1.95(0.95-3.98) | 0.068 |  |  |
| **First marriage and first birth interval (months)** | 1.00 (0.99-1.00) | 0.548 | 1.00(0.99-1.00) | 0.429 |  |  |
| **Gender of index child** |  |  |  |  |  |  |
| Female | 1 | . | 1 | . | 1 | . |
| Male | 0.84 (0.73-0.97) | 0.016 | 0.79(0.68-0.91) | 0.002 | 0.79 (0.68-0.92) | 0.002 |
| **Wealth quintiles** |  |  |  |  |  |  |
| Two poorest | 1 | . | 1 | . |  |  |
| Middle | 0.81 (0.69-0.95) | 0.010 | 0.89(0.75-1.07) | 0.210 |  |  |
| Two richest | 0.76 (0.65-0.89) | 0.001 | 0.91(0.75-1.11) | 0.371 |  |  |
| **Ethnicity** |  |  |  |  |  |  |
| Urdu | 1 | . | 1 | . |  |  |
| Sindhi | 1.39 (1.1-1.75) | 0.005 | 1.05(0.79-1.40) | 0.747 |  |  |
| Punjabi | 1.07 (0.88-1.32) | 0.506 | 1.07(0.86-1.33) | 0.529 |  |  |
| Other | 1.23 (1.06-1.42) | 0.007 | 0.97(0.81-1.16) | 0.745 |  |  |

A- Bivariate analysis
B- Model includes all predictors regardless of their significance in bivariate analysis
C- The predictors significant at $p < 0.2$ in bivariate analysis considered for adjustment. Parsimonious model selected using backward elimination, $p$ -value $< 0.05$ considered significant

Two multivariate models were generated, with model B adjusted for all variables and model C adjusted for significant explanatory variables (Table 3). When adjusted for all explanatory variables, women who were less than 20 years of age were more likely and those between the ages of 25-29 years and 30+ years were less likely to have SBIs compared to women 20-24 years of age. Similarly, women with secondary, and intermediate education also had fewer SBIs compared to those with no education. Couples who did not use contraception within nine months of the index birth, and women who were between 25-29 years at first birth were more likely to have SBIs and those with more male children were less likely to have shorter birth intervals.

Mother’s age, mother’s education, contraceptive use within 9 months of index birth, age at first marriage, and gender of child born prior to index birth were fitted into a Cox proportional hazards model (Model C) and were found to be significantly associated with SBIs. Similar to model A, women between the ages of 25-30 years (aHR=0.64, 95% CI: 0.54-0.77) and 30+ years (aHR=0.30, 95% CI: 0.23-0.40) were less likely to have SBIs compared to women 20-24 years of age; women who had attained secondary (aHR=0.78, 95% CI: 0.65-0.93), intermediate (aHR=0.63, 95% CI: 0.49-0.82), and higher education (aHR=0.71, 95% CI: 0.53-0.96) had fewer SBIs compared to those with no education; and having a male index child resulted in SBIs (aHR=0.79, 95% CI: 0. 68-0.92). Moreover, women who were less than 20 years of age (aHR=1.32, 95% CI: 1.03 to 1.70) compared to women 20-24 years, women who were 25-29 years of age (aHR=1.55, 95% CI: 1.18-2.03) compared to women 10-19 years, couples who did not use contraception within nine months of the index birth had a higher likelihood for SBIs compared to those who used contraception (aHR=2.33, 95% CI: 2.01-2.70). Kaplan-Meier survival curves depict the probability of SBIs by the various subgroups (Figure 1).

**Figure 1:**
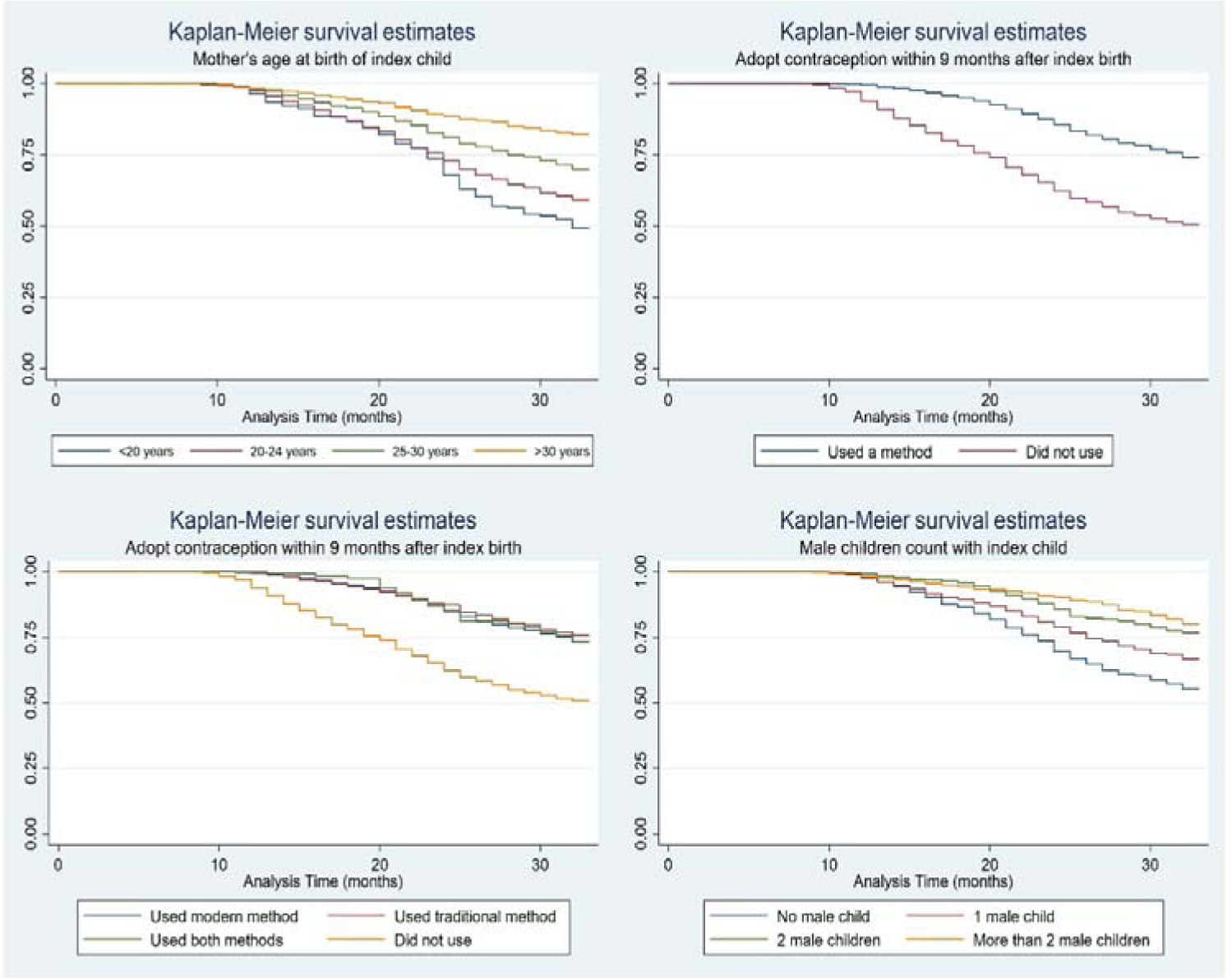
kaplan meier survival estimates.

## Discussion

Short birth intervals are associated with adverse neonatal outcomes and neonatal mortality; and contribute to the burden of disease among neonates in LMICs.^43^ This study evaluated the predictors of SBIs (<33 months) in urban areas of Karachi, Pakistan, and found that 22.9% of births that occurred within six years of the study had a following birth less than 33 months of the index birth. The average median birth interval in our study was 25 months, which is lower than the national median of 29.8 months in urban areas, and lower than the recommended duration.^35^ This interval is much shorter than study conducted in neighbouring Iran where the median duration between two live births was 39 months,^40^ but higher than a study in rural Uganda where the median birth interval was 22 months.^44^ Another large-scale cross-sectional study in rural Bangladesh found that 24.6% had SBIs of less than 33 months, which is very similar to our results, although our study was conducted in an urban setting.^45^ The median birth interval duration reported in our study is also relatively less than the ones obtained in similar studies conducted in Ethiopia,^46^ Myanmar,^47^ and India^48^ with values ranging between 30 to 32.6 months.

In our study, maternal age, education, contraceptive use within nine months of the index birth, and gender of the child prior to the index birth were the strongest predictors of SBIs. Maternal age was a major determinant of all birth intervals in a similar study in Pakistan on the determinants of higher-order birth intervals, where increasing maternal age increased birth intervals.^49^ These findings are also consistent with those reported from Bangladesh, where mother’s age at first birth, parity, survival status of the index child, mother’s education, place of residence, and family composition i.e. having a male child was significantly associated with length of birth intervals.^50^ Similarly, in Iran, the current age of women and maternal age at the time of delivery were strongly associated with birth interval duration.^45,51^ Our results correspond with a study in Uganda where SBIs were associated with younger maternal age.^44,45^ The Pakistan Demographic Health Survey (PDHS) 2017-18 also found that younger women had SBIs compared to older women.^35^ This could possibly be due to the increasing maternal age not only raises concerns for infertility; but also motivate woman to quickly have her desired number of children. In addition, women have more autonomy in making reproductive decisions when they are older.^44^ Moreover, older women are also more likely to have achieved their desired family size and therefore have longer birth intervals.^46^

As expected, women who did not use any contraceptive method nine months prior to the index birth were also more likely to have SBIs compared to those who used any form of contraception. The results are consistent with the findings from a literature review of 14 studies conducted in developed and developing countries which found the use of contraceptive is protective against SBIs.^52^ Though, many of the published evidence in this domain from Pakistan is 20 years old^53-55^ studies from Bangladesh has and India supported the evidence.^48,50^ Similar findings have been reported from Africa, where lack of contraceptive use was found to be one of the strongest predictors of SBIs in Ethiopia.^56^ We found that women with higher education were less likely to have SBIs, which is in concordance with studies from Bangladesh and Saudi Arabia.^45,50,57^ A study in India found that education and women’s autonomy were both strongly associated with longer birth intervals.^58^ An analysis between education and fertility in Indonesia proposed that women who are more educated have a higher likelihood of participating in family planning programs, using services and using long-acting modern contraceptives since they have more knowledge of birth control methods and utilize them accurately.^59^ Moreover, educated women are likely to marry later and thus limit their reproductive years and number of children.

Another finding of our study was that women who had a male index child had a reduced likelihood of SBIs than those who gave birth to a female child. Parental attitudes and preference for male children in Middle Eastern and South Asian cultures may be the reason for this finding, since male children are typically regarded as economic assets as well as future bread earners for the family.^60–62^ Societal pressures for a woman to demonstrate her fertility and for her to bear a son may be influencing her ability to make decision around the spacing of children and use of contraceptives ^60,63^ ^ _recen_j- study was conducted analyses using three DHS from Pakistan on preferences for male children and its impact on birth intervals. They found a significant impact of son preference on birth intervals during the first two parities, where women who had daughters had significantly shorter subsequent birth intervals compared to those who had more sons.^39^ Moreover, women with one or more sons were more likely to use contraceptive methods, indicating a strong preference for sons compared to daughters.^64^ In order to tackle this pervasive desire for male children, gender equality measures, importance of girls, and awareness is crucial. This has major policy implications for the family planning programmes which should be questioned for investing more money into motivational campaigns and should have more integrative policies to promote education for girl child, implementation of legislation against discrimination on the grounds of sex, abolition of practices such as dowry and bride prices, and promoting social welfare and social security so a son is no longer considered an asset and security for an old age.

### Strengths and limitations

This is one of the first studies to investigate birth spacing in urban areas of Karachi, Pakistan. The study, however, is not without limitations. First, our study was conducted in selected low-income areas of Karachi, Pakistan, and is therefore not representative of the national or the local population. Second, our analyses do not include pregnancies that resulted in abortions or miscarriages, and therefore, when live or still-births are preceded by a non-live pregnancy, there could have been an underestimation of the proportion of closely spaced pregnancies. Third, the determinants identified are only for births that occurred within our study period, and it is possible that other variables could have played a role in predicting birth intervals in the participants in our study. Finally, due to the six-year time frame, children born to women in our study before or after the time period were not included and were therefore counted as no-event. The last-born infant of each woman in the study timeframe was also included as no-event since there was no data for live births after that infant, and this may have introduced an under-representation of the number of SBIs in our study.

## Conclusion

Optimal birth spacing has the potential to improve maternal, neonatal and infant health outcomes, reduce familial financial burdens, and allow parents to provide children with comprehensive care and attention. Our findings suggest that reproductive health interventions should address underlying socioeconomic factors that contribute to SBIs, such as preferences for male child, education, and younger MRWA. Family planning should be integrated with other multi-sectoral programs such as education, where girls from the early stage should be empowered and given awareness on these issues. Moreover, family planning strategies should not only focus on increasing coverage of services but also to create awareness about optimal birth intervals and interventions to enhance modern contraceptive utilization behaviours among women of reproductive age.

## Data Availability

The data of the study is available upon request keeping in view institutional and ethical polices of the Aga Khan university by emailing to corresponding author.

## Acknowledgements

We would like to thank all study participants, including surveyors, data enumerators and team leads.

## Author Contributions

SBS was the principal investigator for Pakistan. SBS. IS, DC conceptualized the idea & designed the study. SN, MB & ZS drafted the manuscript. AR and UA managed and analysed the data. IH & KH were involved in the implementation of project and contributed to the development of monthly, quarterly and final report and manuscript. SBS, DC, KH & SI interpreted the data and critically reviewed the manuscript. All authors contributed in manuscript review and approved the final manuscript.

